# Building Resilient and Inclusive Primary Health Care Systems to Improve Access and Vaccine Uptake During a Pandemic: A Systems Thinking Analysis Using Group Model Building for Persons with Disabilities

**DOI:** 10.64898/2026.05.11.26352873

**Authors:** Allan Mayaba Mwiinde, Isaac Fwemba, Patrick Kaonga, Joseph Mumba Zulu

**Affiliations:** Department of Epidemiology and Biostatistics, School of Public Health, University of Zambia, Lusaka, Zambia; Department of Medical Education, School of Medicine, University of Zambia, Lusaka, Zambia; Department of Health Promotion, School of Public Health, University of Zambia, Lusaka, Zambia; Department of Health Policy and Management, School of Public Health, Lusaka, University of Zambia, Lusaka, Zambia

**Keywords:** Primary Health Care, Resilience, Inclusive, Vaccine Uptake, Persons with Disabilities, Health Systems

## Abstract

Strengthening the resilience and inclusivity of primary health care (PHC) systems during crises is critical to achieving equitable access to health care in low-income countries. The COVID-19 pandemic exposed significant weaknesses in PHC systems, highlighting gaps in inclusivity and resilience, particularly for persons with disabilities (PWDs). Although studies have examined PWDs, few have applied systems thinking approaches such as Group Model Building (GMB). This study aimed to develop a resilient and inclusive PHC system to improve access to services and vaccine uptake among PWDs during pandemics. A mixed-methods design incorporating GMB was employed in three phases. First, quantitative and qualitative data were collected to identify barriers and facilitators to PHC access and vaccine uptake. Second, a stakeholder GMB workshop was conducted in Monze to map system dynamics and develop causal loop diagrams. Third, validation and refinement meetings were held, including a final workshop in Lusaka. Findings identified key endogenous system drivers influencing vaccine uptake and access to PHC services and consumables, including financing mechanisms, human resources, outreach services, transport, staff commitment, and availability of accessible information such as Braille materials. These interact through reinforcing and balancing feedback mechanisms. In addition, critical contextual (exogenous) drivers such as political will, health insurance, community gatekeepers, and road networks shape system performance and influence access and service delivery. Strengthening both endogenous system drivers and contextual factors through inclusive strategies, coordinated financing, and supportive governance is essential for building resilient PHC systems that improve equitable access and vaccine uptake among PWDs during health crises. These findings contribute to Universal Health Coverage and equity by showing that strengthening both health systems drivers and contextual drivers is essential to ensure inclusive, accessible, and fair delivery of PHC services for all populations, including persons with disabilities.

## Introduction

Building a resilient Primary Health Care (PHC) system that can respond to health threats and shocks while maintaining essential health service delivery is one of the critical needs globally [1]. The global public health shocks and threats arising from the Ebola virus, the COVID-19 pandemic, and the ongoing everyday stressors of health highlight the vulnerability of health systems across the world [2].

Having resilient PHC systems is crucial in fostering the achievement of Universal Health Coverage (UHC) and health security [3]. Health System Resilience (HSR) is “the ability of a system, community or society exposed to hazards to resist, absorb, accommodate, adapt to, transform and recover from the effects of a hazard in a timely and efficient manner, including through the preservation and restoration of its essential basic structures and functions through risk management” [4].

However, the concept of health system resilience has been critiqued by researchers for its lack of clear definition, the challenges in measuring it, and the absence of clear strategies to support resilience capacities [5].

Despite the criticism, there has been a growing demand in the literature to adopt this idea. The adopted resolution of the World Health Assembly in 2011, on “Strengthening national health emergency and disaster management capacities and resilience of health systems” has now gained global interest with slightly increasing literature on the subject and evidence base for the concept and application of health systems resilience, triggered by recent and current public health emergencies [6].

The vulnerability of the weak health system was only identified in the Low and Low-income countries (LMIC) due to the relationship between poverty and healthcare access [7]. High-income countries had setbacks such as financial constraints during the global recession period, which reduced funding in the healthcare system, overburdening the service delivery system [8].

With the focus on global health matters for the past 10 years, attention has been given to the weaknesses of health care systems in low- and middle-income countries [9]. The greatest weakness was observed in the African region, struggling to adapt from past experiences, causing some systems to continue facing similar problems time and again. In light of this, if there is an increased focus on learning from crises, this will most likely result in a better understanding of resilience strategies and what works better for the developing nations [10].

For African countries to develop resilient strategies, there is a need to ensure countries can effectively prevent, prepare for, detect, adapt to, respond to, and recover from public health threats while ensuring the maintenance of quality essential and routine health services in all contexts, including in fragile, conflict and violence settings [11].

Due to increased recognition among global stakeholders of understanding the complex adaptive nature of health systems, there are challenges to its low use in developing countries to guide current and new policy development [12]. The range of the interacting systems and influences operating at different levels requires the use of analytical tools and frame work, such as systems dynamic modelling, suited to the analysis of complex adaptive systems, which do not take a linear approach [13].

GMB is increasingly being used to inform decision-making however, there is limited research on how effectively group model building has led to the adoption of evidence-based practices, interventions, and public health policies. The use of systems models can help researchers and policymakers in LMIC anticipate the potential intended or unintended effects of policies, interventions, or changes in context over time on the delivery of Primary healthcare services [14].

One of the important systems dynamic research methods is Group model building (GMB), which is a participatory research method that provides new insight into the etiology of complex problems in the area of scientific domains [15]. GMB is the process of engaging stakeholders in a participatory modelling process to elicit their perceptions and views of a problem to help explore concepts regarding the origin, contributing factors, and potential solutions or interventions to a complex issue, which can be used to guide policymakers for effective decision-making making [16].

Studies have established that though PWDs have access to primary health care services, there is still some limitations in access of primary health care services, such as transportation, architectural, and communication barriers, which have been highlighted in Brazil [17].

It will be difficult to achieve Universal health coverage without providing the much-needed attention to people with disabilities [18]. Creating a resilient Primary Health Care System will not only help in archiving universal health coverage for persons with disabilities but also help to have an improved community healthcare system [19].

Given the high levels of disabilities in Zambia, nearly 11% of the population [20]. It is critical to develop a resilient Primary Health Care system that can absorb shocks while being inclusive of PWDs. Disability inclusion is critical to achieving the Sustainable Development Goals and global health priorities, which aim to achieve health for all without leaving anyone behind.

Therefore, this study aimed to develop a resilient and inclusive primary health care system that improves access and enhances vaccine uptake during pandemics, with a focus on persons with disabilities, using a group model-building approach.

### Methodology

#### Ethics statement

Ethical approval for this study was obtained and granted by the Excellence in Research Ethics and Science (ERES) Converge, reference number 2023-Jan-005. Permission was obtained from the National Health Research Authority (NHRA), Reference Number NHRA0003/05/04/2023. Clearance to collect data was also obtained from the Ministry of Community Development and Social Welfare and the management of the selected districts. All participants provided written informed consent prior to participation, and their confidentiality and privacy were rigorously maintained throughout the study. Parental or guardian written informed consent was obtained for participants with severe disabilities who were unable to provide consent independently. No minors were enrolled in the study. Participants were informed that they were free to stop or refuse to participate in the study at any time without any consequences.

The study was structured according to three distinct phases of activity, following the conventions of systems thinking analysis and adopting a group model-building approach [13]. Phase one involves collecting primary data from 985 participants and conducting 7 focus group discussions across the Three Districts (Lusaka, Mazabuka, and Monze) from 5^th^ June to 31^st^ August 2023. (S1 Appendix, S2 Appendix, S3 Appendix and S4 Appendix). Participant diversity was purposively included to ensure representation of multiple lived experiences, particularly among persons with disabilities, thereby enhancing the richness and credibility of the causal loop diagrams development process with the identified stakeholders.

This manuscript shares the same data set as the published manuscript by Mwiinde et al., [21].

#### Study setting

The three Districts were sampled at random to represent settings with a varied fragility profile and diversity among PWDs. The capital of Zambia is Lusaka, which has the largest population of PWDs. Mazabuka and Monze encompass rural and urban township areas, offering information from both urban and rural contexts.

#### Group Model Building

The study used Group Model Building (GMB) to understand the operationalisation implementation strategies for improving vaccination and access to PHC among PWDS. This is because this method supports evidence implementation in real-world practice and describes how GMB can aid in selecting and tailoring both health interventions and implementation strategies that are accepted and appropriate for a local context and acceptable to decision-makers and community members [22]. The participatory epidemiological approach was used to collect data from questionnaires and focus group discussions (S2 Appendix and S4 Appendix), which were to be validated and approved by stakeholders with experience in the PHC systems.

#### Phase One: Planning for the GMB session: The Case of Mazabuka District

The modelling team conducted 4 meetings in one month in order to identify the stakeholders involved in the vaccination and PHC systems at the district level, and develop the agenda for the GMB session [23]. The GMB team identified and purchased the required tools to be used for the GMB session. The identified stakeholders were written to with letters of invitation to participate in the GMB session. All stakeholders who were invited accepted to attend the session.

Study participants were purposively sampled from stakeholders involved in community leadership, community health volunteers, Ward Development Committees (WDCs) and non-governmental organisations involved in working with PWDs, and government health workers involved in PHC services at the district level. A variety of data collection activities were undertaken, using participatory epidemiological discussions with identified key stakeholders (actors) of 20 participants from Mazabuka. The stakeholder mapping process used a participatory method through a matrix (Fig 1), which uses the power and level of interest of a stakeholder involved in disease prevention, working with vulnerable groups in improving the health outcomes (Thompson, n.d).

**Figure 1:**
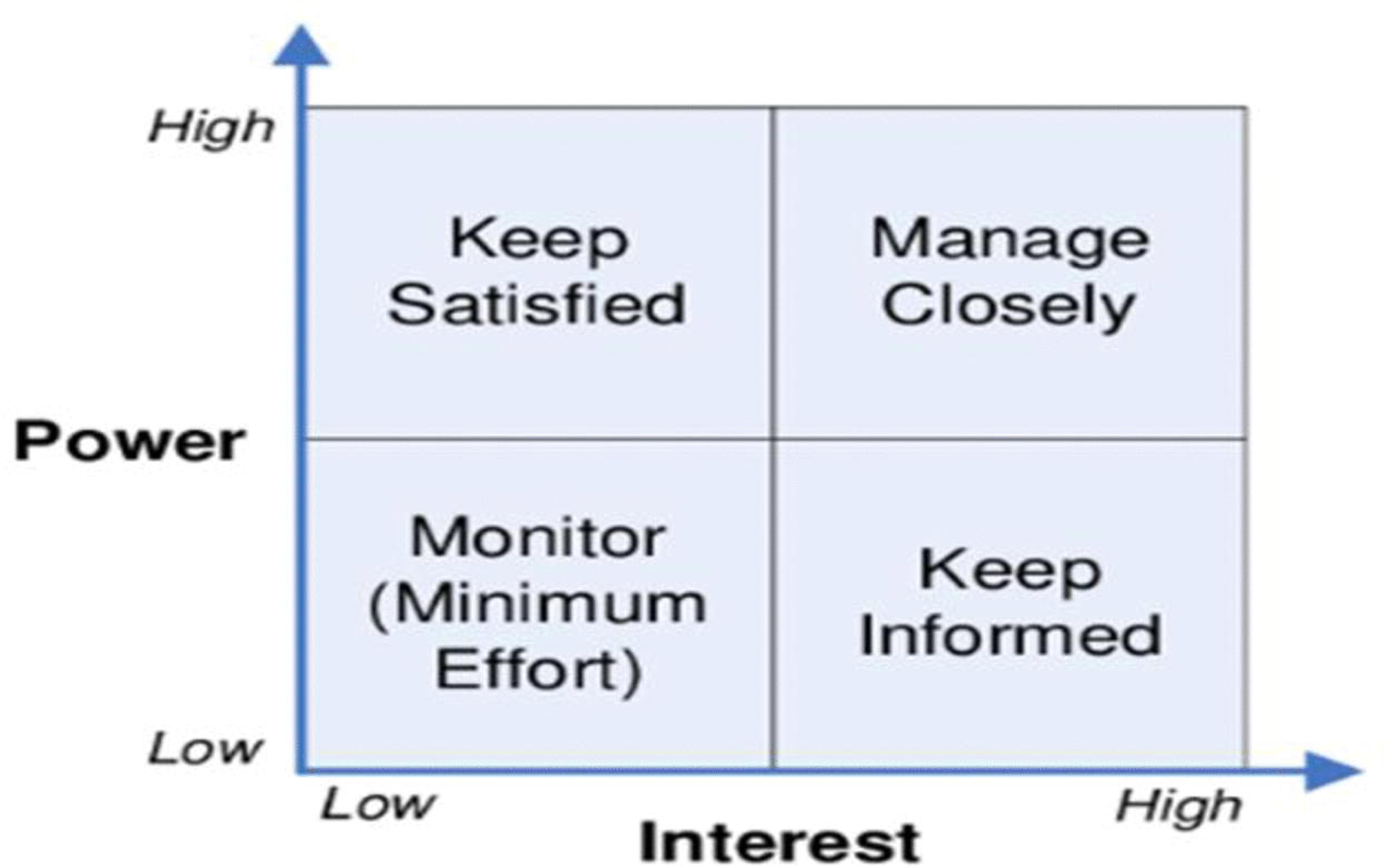
This is the figure 1 Title. Stakeholder Mapping process for participation in Group Model Workshop.

To identify the general themes to be discussed in the GMB workshops, the research team met and preliminarily analysed the results of a structured questionnaire and focus group discussions, to identify prevalent themes and those that are key in improving PHC at the community level to be explored in the GMB workshop based on the obtained data (S 2 Appendix and S3 Appendix) [23].

Group model building: data collection and analysis were presented at the GMB workshop using the methodological tool used by systems thinking researchers, which allows researchers and relevant health stakeholders to come together and, in a participatory manner, elaborate conceptual models of system behaviours/problems under study. During this study, only one GMB workshop was conducted with 21 participants with an interest in PHC. The compositions of participants were from the District’s Health Office (Human resource, vaccination program officers, Environmental Health Officer, Health promotions officer, District Health Officer, Nurses in Charge, pharmacist in charge, and accountant) Mazabuka Municipality (Public Health and Engineering Department), The Ministry of Social Welfare, the Ministry of Community Development the Community Healthcare workers, and persons acting as caregivers to the PWDs.

#### Phase Two: GMB venue and assignment of responsibilities

Day one: The GMB team meet at the venue of convening. The team organised the venue and the seating arrangements during the GMB session. The team shared the tasks to be conducted during the GMB session and developed the schedule of activities according to the agenda.

Second day. The GMB sessions were organised around a series of scripts that correspond to sequential activities carried out by the participants, which are used in the study. Scripts detailing each activity were elaborated and used by all workshop facilitators to ensure consistency in prompts given and activities undertaken by the diverse participant groups. The GMB session consisted of three sections

#### Phase Three: GMB Workshop

Participants were introduced to Group Model Building (GMB) sessions and the rationale behind them as facilitated, structured workshops designed to elicit participants’ expertise and translate it into visual system models across several key stages. These stages were clearly explained to participants, including: introduction and problem identification; variable elicitation; development of causal loop diagrams; identification of leverage points; prioritisation; and action planning.

#### Script 1: Personal Introductions, Hopes and Fears of the GMB team

The GMB session started with the stakeholder’s individual introduction with key introduction words such as the name of the participants, where they are coming from and what they do and like doing the most during their free time. In the introduction, the team agreed to use appropriate words such as PWDs instead of using the word such as “disabled people” which is discriminatory. The second questions requested the participants to mention their hopes and fears. On a sticker of the same colour which were given to participants. The GMB recognizes that the quality and validity of model outputs are contingent on the level of trust, openness, and inclusivity within the group process [24,25]. The “Hopes and Fears” exercise served as a critical entry point for surfacing participant expectations, concerns, and potential power dynamics, thereby strengthening psychological safety and promoting equitable participation. Addressing these factors at the outset enhances methodological rigor by reducing the risk of dominance bias, social desirability bias, and constrained participation—common threats to validity in group-based qualitative approaches.

#### Script 2: Visualizing barriers to vaccination and accessing PHC among PWDs

Building on this foundation of the first script, the second script was designed to facilitate discussion on key barriers to vaccination and access to PHC among PWDs. Participants were invited to construct Rich Pictures to visualise the existing system, enabling them to identify and articulate critical challenges affecting access to vaccination and PHC services within their communities and across the district. This progression from establishing shared understanding to problem structuring supported the development of more comprehensive and contextually grounded system insights. [26].

#### Script 3: System Conceptualisation and Identification of Fragility and Intervention Points

Third, participants were asked to review the rich pictures developed and abstract a key set of variables relating to the onset of COVID-19 vaccine programmes, and access to primary healthcare services among PWDs. Using this concept, researchers assisted participants in building a preliminary concept model describing the dynamics of PHC in the context of a pandemic, such as COVID-19, in accessing vaccines and PHC services. Fourth, participants were prompted to identify particular points of fragility (i.e. areas of particular weakness) and also intervention (i.e. areas where intervention would be most strategic) in the depicted concept [27].

#### Script 4: Causal Loop Diagram Development and Identification of Leverage Points from Participant Models

Connections between the variables as described by participants in their initial concept models were translated into an electronic model using Stella Professional Version 3.7.2 [28]. Similar to interviews, GMB models underwent iterative analyses: first, using notes from the GMB sessions, variables and pathways in the concept models were refined and as needed consolidated to ensure the concept models reflected in the underlying causal logic of participants. The resulting causal loop diagrams then underwent further iterative critical analyses. This entailed comparing variables and their definitions, comparing pathways to ensure consistent and divergent information is captured and highlighting pathways and variables specific to only one group of participants. Balancing and reinforcing loops were then identified in this final causal loop model [29]. Further, researchers discussed the leverage points affecting system behaviour and compared these to the points identified by participants as fragility/intervention points. Associated interventions, as elaborated by participants, were then noted alongside the causal loop diagram according to the variables affected.

GMB further focused on reviewing and refining the initial system map from the participants’ group to ensure that there was consensus and participants’ views and experiences were captured accurately. Model updates were captured in STICKE in real time. Achievable action ideas, including who should be a part of intervention design were identified during the workshop. The facilitators chose not to interrupt these conversations, ensuring that every participant had the opportunity to be heard.

### Phase Four: Post-GMB Workshops

#### Script 5: Refinement of the Causal Loop Diagrams and Categorising the System-Level Structure

Two post–Group Model Building (GMB) workshop meetings were conducted. The first meeting involved seven participants from the initial GMB workshop, who were invited to review the final model output and suggest any necessary revisions; only minor refinements were made.

The second meeting, held in Lusaka, involved a presentation of the GMB findings to a broader group of stakeholders. This group included seven academics, three of whom had experience in implementation science research within PHC systems. The remaining ten participants had experience working in primary healthcare (PHC) systems, and all were researchers with a focus on PHC. This session led to further refinement of the model, particularly in identifying actionable variables likely to influence PHC. Following the final development of the causal loop diagram, endogenous system drivers were systematically categorised into three interrelated domains to support analysis and interpretation of the model. This categorisation was used to organise variables within the model and to facilitate examination of their interactions within reinforcing and balancing feedback loops influencing access to PHC services and vaccine uptake. Exogenous system drivers were identified during the post-model refinement phase through further stakeholder engagement and were incorporated to ensure the model adequately reflects external contextual influences [30]. This stage marked the final validation and approval of the causal loop diagram (CLD).

## Results

Causal loop diagrams (CLDs) were developed by the GMB facilitation team based on key system variables identified from a secondary dataset (N = 985) (Table 1). These variables informed the initial system structure, which was subsequently validated and refined through stakeholder input during Group Model Building workshops.

**Table 1:** Demographic Variables and their Contribution to Group Model Building.

| Variable | Variable Description | N = 985 (%) | Contribution to GMB |
| --- | --- | --- | --- |
| Age | Age category |  | Ensured representation of diverse life-course experiences influencing health service access and vaccine uptake perspectives. |
|  | 18–25 | 205 (20%) |  |
|  | 26–35 | 168 (17%) |  |
|  | 36–55 | 289 (29%) |  |
|  | 56+ | 323 (33%) |  |
| Sex | Male | 489 (50.4%) | Enabled inclusion of gendered experiences in accessing primary health care and vaccination services. |
|  | Female | 496 (49.6%) |  |
| Marital Status | Marital Categories |  | Provided insights into household support systems and decision-making dynamics affecting health-seeking behaviour. |
|  | Single | 435 (44%) |  |
|  | Married | 286 (29%) |  |
|  | Divorced | 78 (7.9%) |  |
|  | Widowed | 186 (19%) |  |
| **Districts** | District Categories |  | Captured contextual and geographic differences in access barriers across urban and peri-urban settings |
|  | Lusaka | 543 (55%) |  |
|  | Mazabuka | 205 (21%) |  |
|  | Monze | 237 (24%) |  |
| **Setting** | Urban | 869 (88.22%) | Informed understanding of spatial accessibility, transport barriers, and service availability in PHC systems |
|  | Rural | 116 (11.78%) |  |
| **Education** | Education Categories |  | Influenced variation in health literacy, understanding of vaccination messages, and system navigation capacity. |
|  | No school | 137 (14%) |  |
|  | Primary | 441 (45%) |  |
|  | Secondary | 362 (37%) |  |
|  | Tertiary | 45 (4.6%) |  |
| **Disability type** |  |  | Central to identifying accessibility barriers, communication challenges, and structural exclusion within PHC and vaccination systems. |
|  | mobility impairment | 730 (74%) |  |
|  | Spinal cord disability | 205 (21%) |  |
|  | Traumatic brain injury | 146 (15%) |  |
|  | visual impairment | 247 (25%) |  |
|  | speech disability | 177 (18%) |  |
|  | cognitive disability | 174 (18%) |  |
|  | psychological disorders | 152 (15%) |  |

1. Disability Type: Multiple responses were allowed for each individual; therefore, percentages do not sum to 100%.

The secondary dataset showed that the largest proportion of respondents were aged 56 years and above. The sex distribution was nearly equal, with females accounting for 49.6% and males 50.4%. In terms of marital status, single participants represented the largest group (44%). Geographically, the majority of respondents were from Lusaka District, and most participants (88%) resided in urban settings. Regarding educational attainment, primary education was the most common level (45%). In terms of disability type, mobility impairment was the most frequently reported category (74%) among participants.

### Participatory Group Model Building

The modelling process followed an iterative refinement pathway, progressing from stakeholder-generated representations to a synthesized final causal loop diagram that captures the key system dynamics influencing vaccine uptake and access to PHC services

### Participatory Group Model Building of Rich Pictures

Participatory Group Model Building produced the initial visual representations through which participants conceptualized challenges related to access to vaccines and primary healthcare (PHC) services among persons with disabilities (PWDs) (Figs 2 and 3).

**Fig 2.**
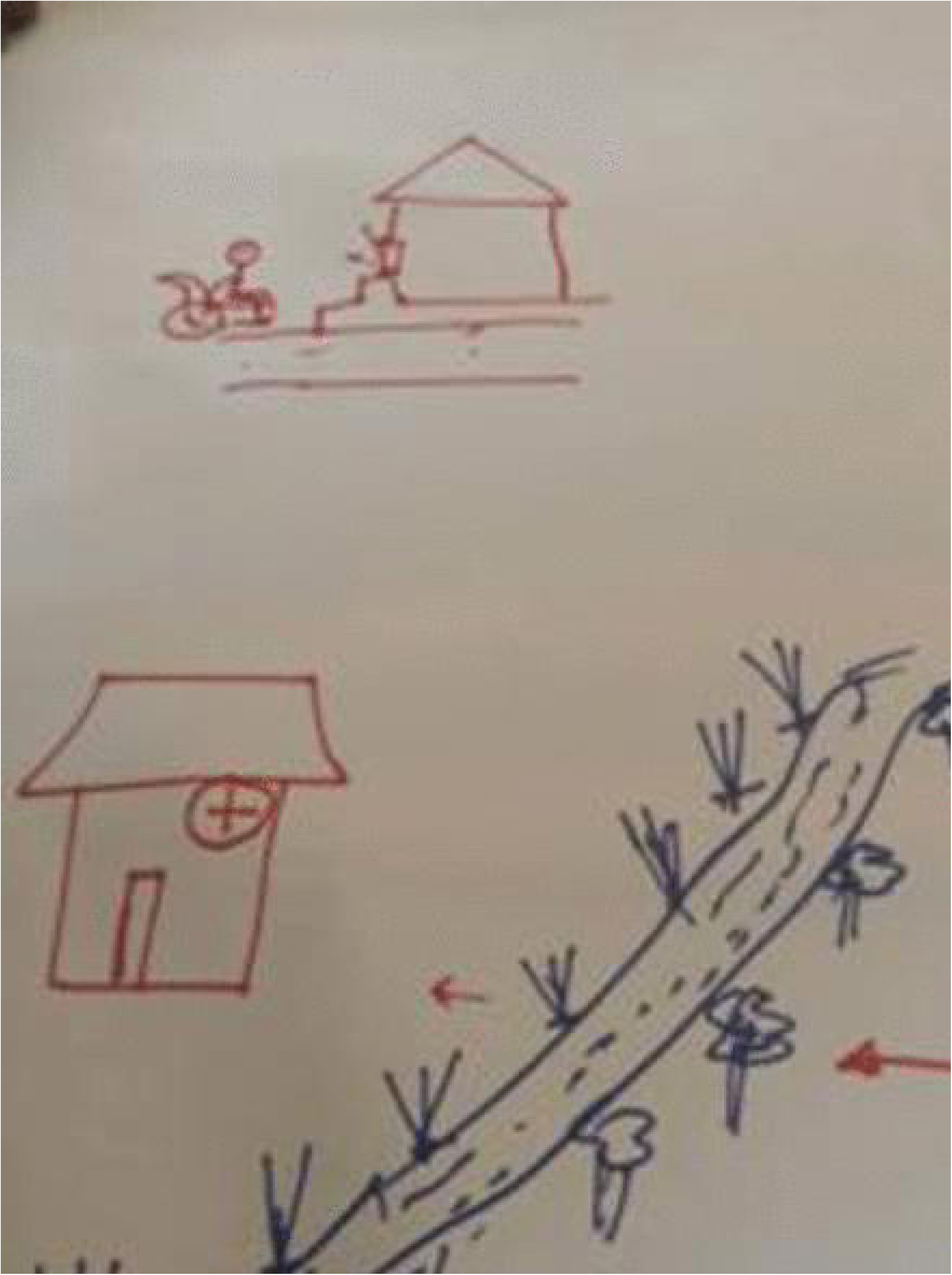
This is the figure 2 and 3 Title. Rich Pictures drawn by the Participants during GMB (i) Movement disability failing to cross the stream (ii) Vision disability being by-passed to the vaccination table.

**Fig 3.**
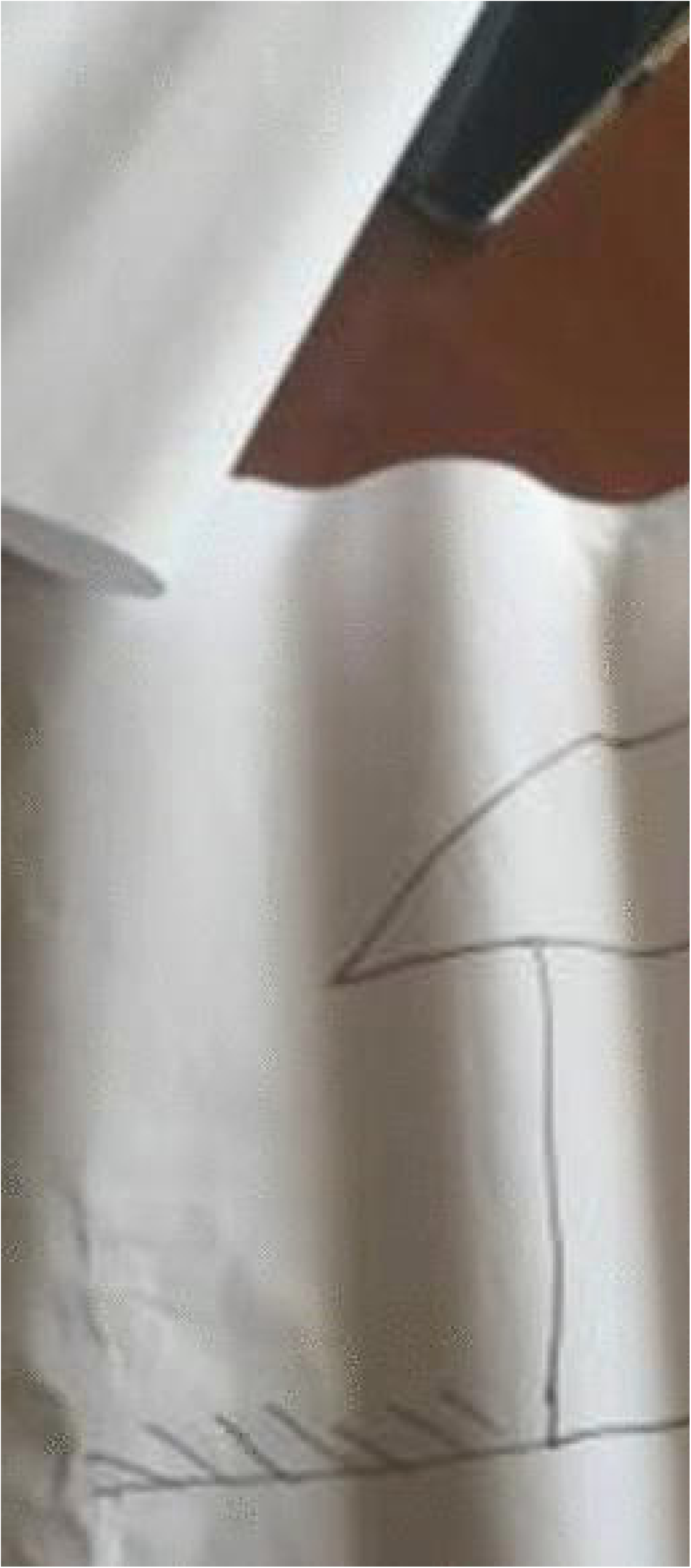
This is the figure 2 and 3 Title. Rich Pictures drawn by the Participants during GMB (i) Movement disability failing to cross the stream (ii) Vision disability being by-passed to the vaccination table.

### Participatory Group Model Building Process Results

#### Understanding Key Variables from the Participatory Group Modell Building

Eighteen variables were confirmed by GMB participants as key to creating a resilient PHC system that can withstand shocks and challenges in an epidemic or pandemic. Seven other factors were identified as intervening factors between vaccination, human resources, and outreach.

The results of the session after cleaning and selecting key variables for the final loop diagram were selected for the final input of the loop diagram of improved uptake of vaccines and access to PHC (Fig: 3). Vaccination and access to healthcare were identified as seed models for the GMB, providing a basic structure on which to build, without unduly influencing model development. Financing was an influencer in that the purchase of the vaccines reduced the finances, while the uptake of vaccines greatly improved the quality of life. Access to PHC reduces finances. However, access to PHC improves the quality of life (Fig 3).

#### Stakeholder-Derived Models: Identification of Key System Variables

The initial stakeholder-derived model captured participants’ perspectives on barriers and enablers influencing vaccine uptake and access to PHC services among people with disabilities. This model highlighted a broad set of interrelated factors, including financing, service availability, health workforce capacity, community engagement, and sociocultural influences. Relationships between variables were largely descriptive and non-directional, reflecting the exploratory nature of this stage. This stage provided a foundational understanding of the system, ensuring that the modelling process was grounded in lived experiences and context-specific realities. Key variables identified here informed subsequent abstraction and structuring of the system. Building on these stakeholder insights, the variables were synthesized into a simplified structure to identify core system dynamics beginning with the first seed model.

The seed model identifies financing as a central driver linking vaccine uptake and access to PHC facilities, operating through two key feedback mechanisms. The reinforcing loop (R1) illustrates a positive feedback process in which increased vaccine uptake can attract or justify additional financing, which in turn supports further improvements in uptake. However, this same loop may also contribute to financing depletion over time, as higher uptake increases demand for vaccines, service delivery, and operational costs, thereby placing sustained pressure on available financial resources (Fig 4).

**Fig 4:**
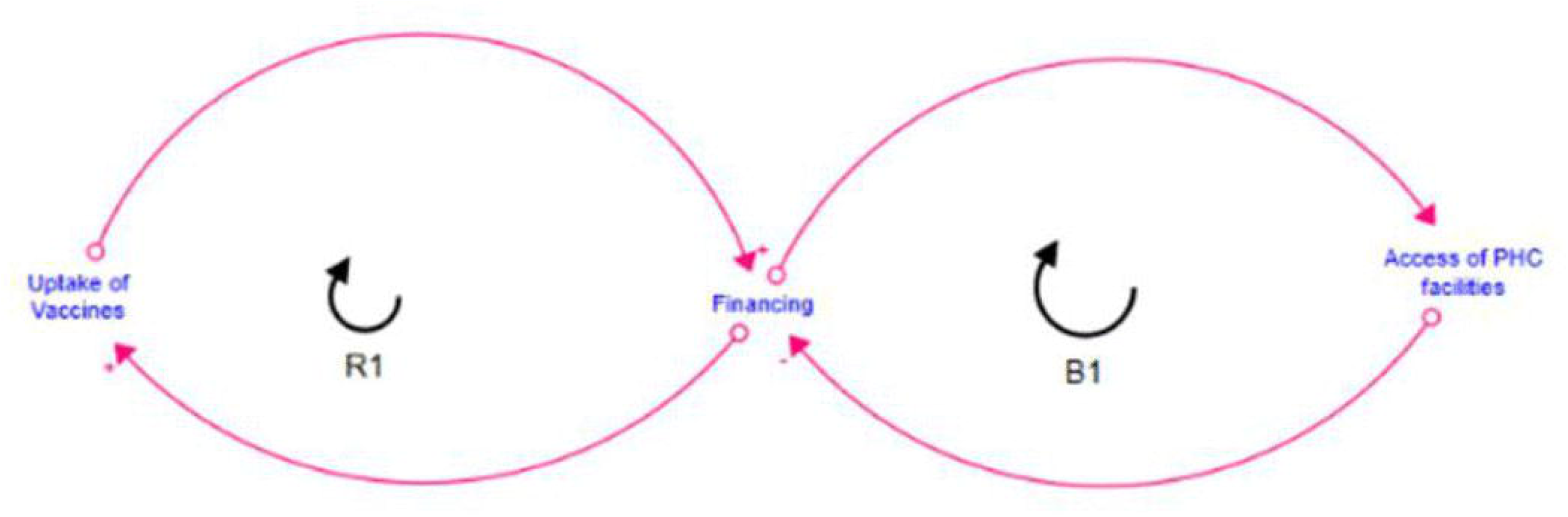
This is the figure 4 Title. Causal loop diagram of seed model on determinants influencing uptake and access of vaccines and PHC among vulnerable groups

The balancing loop (B1) describes the relationship between financing and access to PHC facilities. Increased financing improves access by expanding services, infrastructure, and workforce capacity. Nonetheless, as access improves and utilization increases, the demand for services may exceed available resources, contributing to rising recurrent costs and potential strain on financing. This introduces a stabilizing effect, whereby resource limitations constrain further expansion together, these interacting loops highlight the dual role of financing as both an enabler and a limiting factor within the system. While increased investment can drive improvements in vaccine uptake and access to PHC services, sustained gains are contingent on the capacity of the system to manage rising demand and prevent the depletion of financial resources (Fig 4).

#### Final Model: Consolidation of System Dynamics

The final model integrated stakeholder inputs and seed model structures into a coherent causal loop diagram with clearly defined feedback loops. Reinforcing and balancing loops were explicitly labeled, and relationships between variables were standardized in direction and polarity (Fig 5).

**Fig 5:**
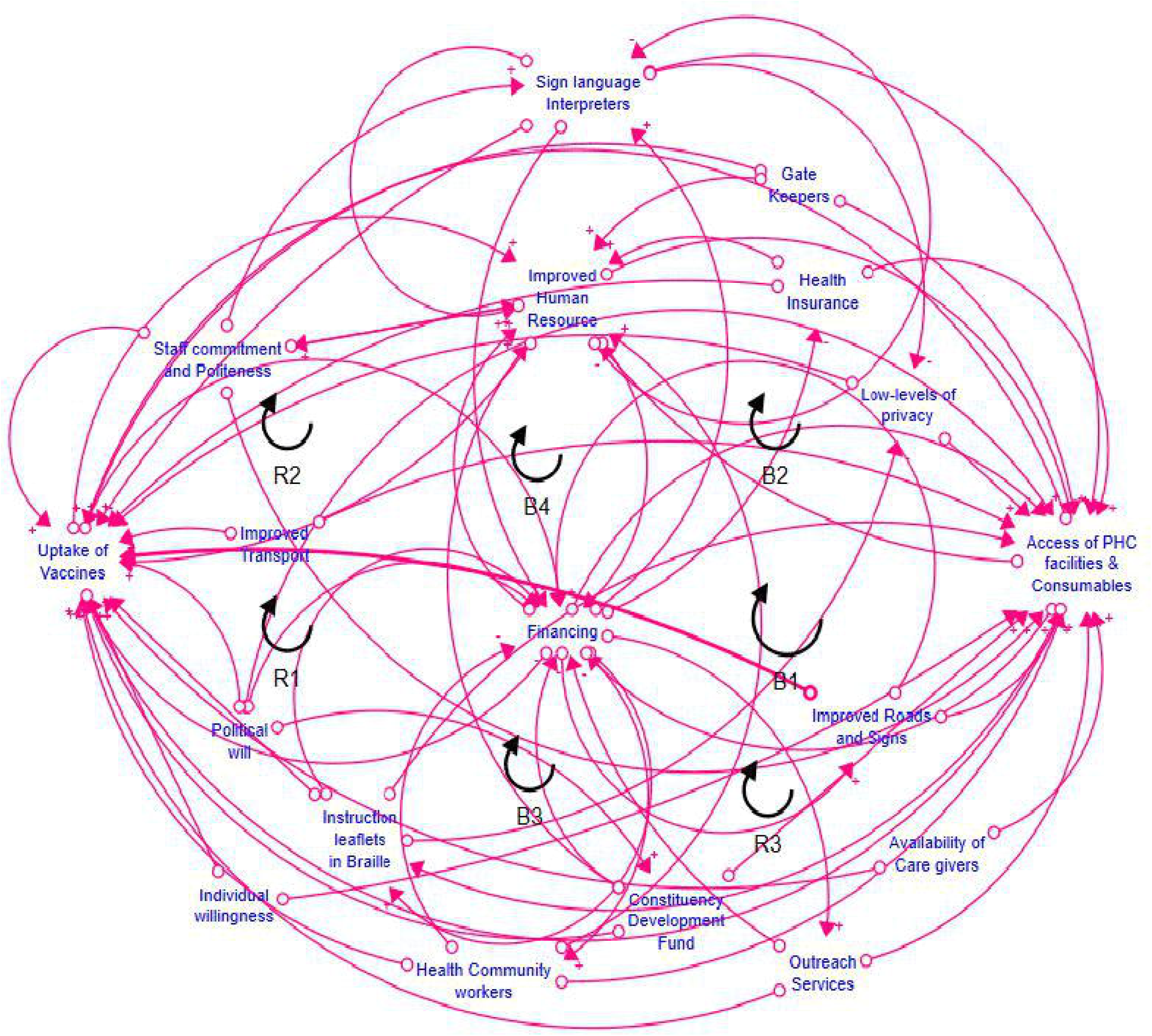
This is the figure 5 Title. Causal loop diagram illustrating factors influencing access to primary healthcare and vaccine uptake among persons with disabilities

The model demonstrated how financing simultaneously drives improvements in vaccine uptake and access to PHC services, while also acting as a limiting factor due to increasing system demands. The reinforcing loop highlighted how increased uptake can stimulate additional investment, whereas the balancing loop captured constraints arising from resource limitations and system capacity (Fig 5). This stage provided a comprehensive representation of the system, enabling interpretation of dynamic interactions between resource allocation, service delivery, and health service utilization. Following validation and feedback from participants, further refinements were made to enhance the representation of key outcomes and system constraints as illustrated (Fig 5).

#### Causal loop diagram analysis of the final Model

The final causal loop diagram (CLD) depicts the interactions between key system variables influencing vaccine uptake among persons with disabilities (PWDs) within primary healthcare (PHC) systems. Financing emerged as a central variable, linking improvements in human resources, access to PHC facilities and consumables, and the provision of inclusive health communication materials (Fig 6).

**Figure 6:**
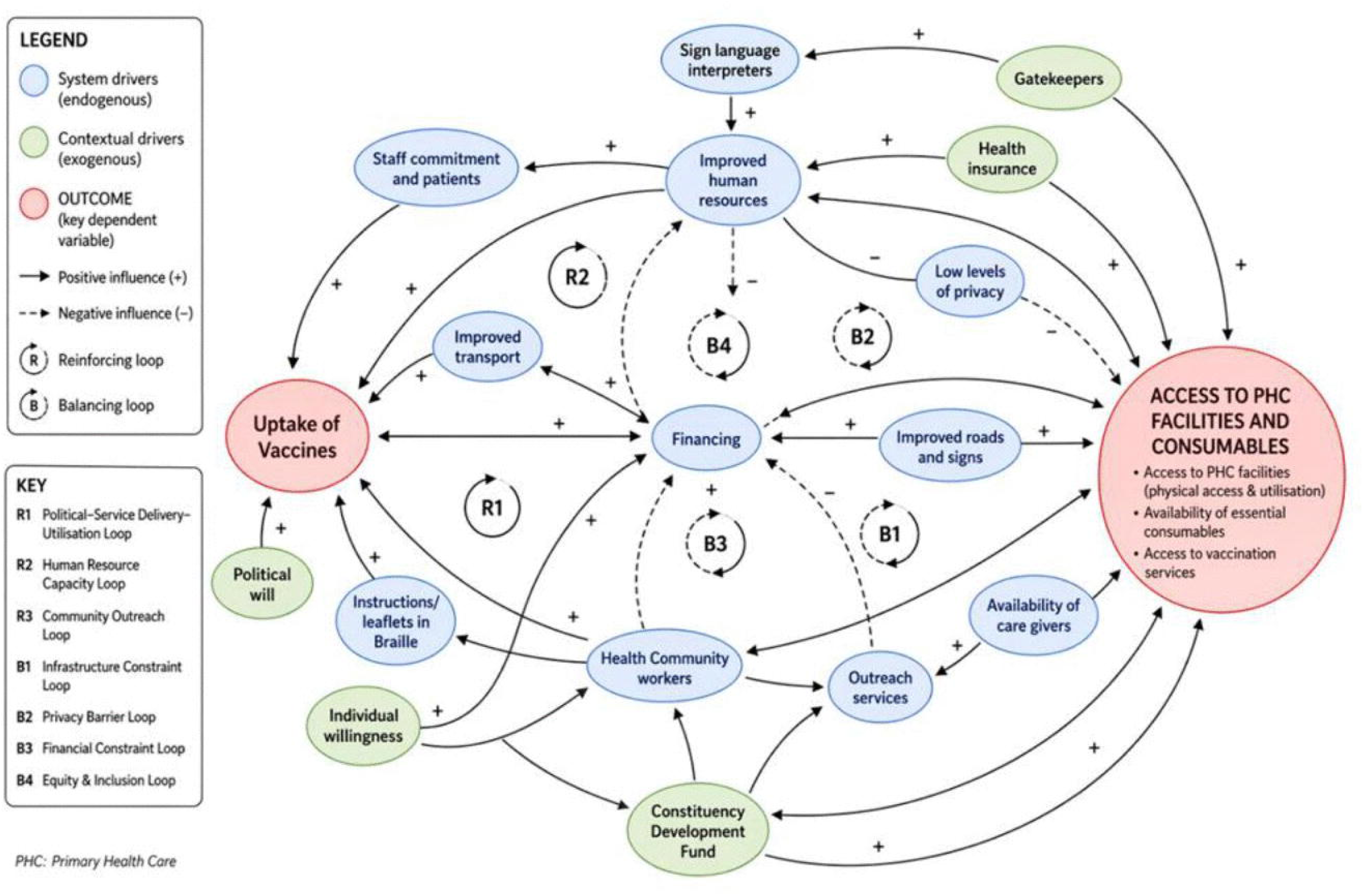
This is the figure 6 Title. Causal loop diagram showing system and contextual drivers influencing access to primary healthcare and vaccine uptake among persons with disabilities

The CLD contains both reinforcing and balancing feedback loops. Three reinforcing loops (R1–R3) describe processes that may increase vaccine uptake over time. In R1, increased financing is associated with improved access to PHC facilities and consumables, which is linked to higher vaccine uptake; increased uptake, in turn, feeds back into financing. In R2, improvements in human resources are linked to enhanced service delivery and increased uptake, which may further support investments in workforce capacity. In R3, the availability of instructional materials in accessible formats, such as Braille, is associated with improved vaccine uptake, which may reinforce the demand for inclusive communication strategies (Fig 6).

Four balancing loops (B1–B4) represent system constraints that may limit these gains. B1 indicates that increased access to PHC services may place pressure on available resources, potentially constraining further improvements. B2 reflects limitations in human resource capacity in response to increased service demand. B3 captures constraints related to the provision and sustainability of accessible communication materials. B4 highlights potential inefficiencies in the allocation and utilisation of financing, which may reduce its effect on other system components (Fig 6).

Comparisons across participant groups identified both convergent and divergent pathways within the model. While the central role of financing and access to PHC services was consistently represented, some pathways particularly those related to inclusive communication were emphasised by specific participant groups (Fig 6).

Leverage points identified within the CLD include financing, human resource capacity, access to PHC services, and inclusive communication strategies. These variables were also aligned with intervention points proposed by participants and were incorporated into the final model as key areas influencing system behaviour (Fig 6).

#### The three System drivers interrelated domains

The final causal loop diagram distinguishes between system drivers embedded within PHC service delivery and contextual drivers operating in the broader environment. System drivers, including financing, human resources, outreach services, and accessibility provisions, actively participate in reinforcing and balancing feedback loops shaping service access and vaccine uptake. In contrast, contextual drivers such as political will, health insurance, and community-level gatekeeping structures influence system performance indirectly by enabling or constraining resource allocation and service utilisation (Fig 6).

System drivers were further grouped into three interrelated domains. First, resource and capacity drivers, including financing, human resources, and availability of caregivers, determine the operational strength of the PHC system. Second, service delivery drivers, such as outreach services, transport, and access to PHC facilities, influence the extent to which services reach target populations. Third, quality and inclusiveness drivers including staff commitment, privacy, and accessibility services (e.g., Braille materials and sign language interpreters) shape the acceptability and equity of service utilisation. These domains interact dynamically within reinforcing and balancing feedback loops to influence vaccine uptake (Fig 6).

### System Drivers of the Causal Loop Diagram

#### Improved Financing in PHC Facilities

Though there are mechanisms to finance primary healthcare facilities already in place in Zambia, there are times when critical need for some essential drugs can only be procured in private pharmacies. This is because some drugs are not stocked in the government facilities, while stockouts are also recorded problems. This means that those who are on long-term medication should be adequately accounted for at their nearest health facilities to identify their medical needs and support. The improved financing of the PHC system will also help to ensure that there are no stockouts of essential drugs in the facilities. This will also improve the vaccine supply chain system by ensuring quality storage, thereby maintaining the cold chain supply systems. One participant in an FGD said, “Health is essential for those who took the COVID-19 vaccine. It was important for prevention to improve life” (FGD 5, Male) (Fig 6).

#### Improved Human Resource

The availability of human resources will help ensure that health services are available at all health facilities and that communities receive vaccination services on time. The inadequacy of human resource personnel can lead to low vaccine uptake and poor access to primary healthcare (Fig 6). GMB Participants indicated that sometimes the queues that are in the health facilities cause challenges to PWDs. Some of them of have mental health problems become inpatients to keep waiting in the queue to seek PHC services. Therefore, having improved human resource personnel in these facilities will reduce the pressure of the long quest for PWDs and the abled population (Fig 6). “*It’s difficult at times to talk about how to reduce the challenges in accessing Vaccines and PHC services because we have talked about this before. However, I said there is a shortage of manpower in these PHC facilities, so if you can increase manpower and see to it that some are specifically to look after the disabled that will improve the situation” (FGD 5, Male)*.

#### Enhanced outreach services

When outreach services are conducted in the communities there is enhanced motivation for PWDs to access the PHC services because they become closer to the doorstep. The highest number of vaccinations during COVID-19 vaccination were during outreach programs. Therefore, outreach programs break barriers of disability once conducted to reach to those who cannot manage to reach the PHC facilities (Fig 6). In a FGD a participants said “Mobile Hospitals and Outreach services would help PWDs with substantial benefits and cushion their movement challenges. In an event of vaccination outreach it is easy to access the vaccine (FGD 5, Male).

#### Availability of instruction leaflet braille’s

There are no reading braille in the PHC facilities for PWDs to be able to read and understand the ongoing pandemic and the available interventions for PWDs to make a decision (Fig 6). In a FGD a participant said “*The availability of the instruction leaf braille to the blind will help them to have firsthand information on health matters happening around. There is also a need to educate the people staying with the deaf and blind, if possible, extend to the community to whoever is willing to learn sign language and large print braille reading to enhance communication among the deaf*” (FGD 4, Male).

#### Understanding of sign language interpreters in PHC facilities

The GMB team understood and discussed that the availability of healthcare workers who can communicate with all groups of people effectively, including vulnerable groups, will create an inclusive healthcare system in which every person can walk in at any time and seek medical healthcare services. This will also promote confidentiality among the PWDs by not having an interpreter as an intermediary with the healthcare worker. This opportunity will also help prevent misdiagnosis in PHC institutions, where sometimes the testing kits and other simple testing types of equipment are not available, and the screening is based on the signs and symptoms of the patients. Furthermore, when there is a direct link between the deaf and the healthcare worker, there is a high certainty of the patients’ understanding that the root cause of the problem has been addressed. The healthcare worker can also easily pass on the necessary health promotion message directly to the patient on how best they can take care of themselves to avoid any infection and worsen the already existing conditions (Fig 6). “*The Government should train health workers to communicate using sign language to end the communication barriers and improve access to PHC among PWDs” (FGD 3, Female). “The Government should employ deaf health workers who can easily understand their language and situation in health facilities to end communication barriers that exists” (FGD 5, Female)*.

#### Staff commitment and kindness

The participants agreed on the need to have healthcare workers who are well-trained to understand the attitudes and perceptions of the PWDs. Some perceptions are created by PWDs that they are not cared for and might need some attention. However, the health personnel must be able to understand and help the PWDs without causing emotional challenges to them (Fig 6).

#### Improved sensitization in communities using instruction reading Braille’s

The availability of communication messages in the communities such as information, education, and communication (IEC) materials that are made into brails for the blind to be able to read and understand some key intervention messages, similar to the way the general population has the opportunity to read the IEC materials, with reduced inequality in access to the health messages among PWDs. In almost all epidemics and pandemics of existing diseases, no IEC material is used to sensitize the PWDs (Fig 6).

#### Good transport system

The availability of transport systems is a challenge for other PWDs who require transportation systems to reach out to the primary healthcare facilities. In some areas, the bus stations are far from the areas where they reside. This makes the PWDs have challenges in accessing the PHC Facilities at a time when they need such services (Fig 6).

#### Improved and Inclusive Health Insurance System

Having access to PHC services by PWDs will help them access the essential drugs that they need from day to day to sustain their living. The PWDs in Zambia are supported through the social cash transfer to help them sustain their livelihood and the public health assistance welfare for medication, which was described as inconsistent for those who qualify to obtain. This amount of Public Health welfare is inadequate and sometimes it is only accessed once a year. However, providing a deliberate policy to include them in the National Health Insurance Scheme (NHIMA) used in Zambia can provide fair access to PHC. Most of the PWDs depend on the government’s PHC facilities. The inclusion of the NHIMA scheme can also enable them to access services at other private facilities at an affordable cost, at a convenient time and place. *“We would like to be on NHIMA at least, that can help us a lot. If the government can help us to have free access to medication like others do, that can improve access to PHC among us PWDs”* (FGD 5, Female) (Fig 6).

#### Accessibility to Infrastructures

Whilst efforts have been made to improve the infrastructure of health facilities accessible to PWDs, not all areas within PHC facilities are accessible. This requires that PWDs always have someone to provide support when visiting PHC facilities (Fig 6).

#### Community Health Promotion

There is a need to enhance community health promotion and participation to improve access to vaccines and PHC services. This ensures that fundamental matters regarding the importance of vaccine acceptance and uptake, and the early seeking of healthcare services, are addressed at the community level. This can reduce the barriers to vaccine uptake and improve vaccination rates. This entails that gatekeepers and community healthcare workers are informed about vaccine development, distribution, and uptake in the community. Community health promotion also enhances individuals’ willingness to receive vaccines and seek primary healthcare (PHC) (Fig 6).

#### Availability of Medical Supplies in PHC Facilities

The availability of medical suppliers in the PHC facility helps enhance access to PHC and improve vaccine uptake. People with disabilities (PWDs) often require repeated access to health services. This indicates a need to improve the stockouts of consumable medical supplies (Fig 6).

### Contextual drivers

#### Good access to the road network system

Most of the PWDs with a physical disability require a good road network system for them to access the PHC facilities easily. However, most PWDs reside in peri-urban and rural areas, where the road network is poor, making it difficult to reach the PHC Facility. Challenges with the road network system include poor gravel roads (potholes and sand), making it difficult to use a wheelchair, flooding of the roads during the rainy season, making them impassible, unmarked tarmacs, and poor usage of the road by motorists without regard for PWDs (Fig. 6).

#### Enhanced Constituency Development funding, Care Givers and Gatekeepers

The funding received by the municipalities from the government, called the Constituency Development Fund (CDF), was identified as a direct contributor to access to PHC. This is because the funding is used to construct health facilities and access roads to these facilities. Caregivers usually help PWDs in accessing PHC and attend to their daily needs. Gatekeepers play a critical role in communities, ensuring that health messages are disseminated to the intended audience (Fig 6).

#### Public Schools for PWDs

The creation of public schools for people with disabilities (PWDs) would not only support their educational needs but also help them read and understand health messages promoted across the country, such as vaccination and personal hygiene messages. People must be made more aware of the need to eradicate stigmatisation across various settings, including families, schools, churches, and public areas. Since PWDs struggle to learn alongside the abled people for reasons that they are stigmatised, the establishment of special schools for them will aid the management of stigma” (FGD 1, Female) (Fig 6).

## Discussion

The study applies GMB to understand structural and interactional barriers affecting PHC access among PWDs, a group often underrepresented in systems modelling research in low-resource contexts such as Zambia. Our findings identified critical variables that contribute to the development of resilient PHC systems, which are essential for mitigating the effects of catastrophic events such as the COVID-19 pandemic[2]. Notably, we highlighted the importance of addressing the needs of vulnerable groups, including persons with disabilities (PWDs). Strengthening PHC to serve these populations effectively is a critical step toward achieving universal health coverage (UHC). This approach enhances PHC system equity, resilience, and overall population health outcomes[31].

Our findings indicate that elements of the endogenous primary healthcare system are likely to meet PWDs’ expectations. Once improved, our findings are similar to those of Harfield et al. [32]: indigenous PHC services are more likely than mainstream services to enhance the health of Indigenous communities they often deliver integrated services that combine treatment, prevention, health promotion, and action on the social determinants of health.

Enhancing health infrastructure and road networks with provisions for persons with disabilities (PWDs) will improve access to primary healthcare services and vaccination during pandemics. Policy measures promoting inclusive planning will enhance vaccination coverage and strengthen primary healthcare delivery for all populations [33]. Our findings follow the recommendation to improve infrastructure to improve the ability to contain the transmission of communicable diseases such as COVID-19 and provide quick access to PHC [34]. Although the health authorities in a decentralised health system have the capacity to detect infrastructural deficiencies and trace the effects of interventions, the road infrastructure is sometimes neglected and not given attention [35,36]. Prioritizing investments in roads and health infrastructure will therefore strengthen system resilience, reduce inequities in access, and improve overall health system responsiveness in times of crisis. Prioritize road infrastructure within decentralized health systems, ensuring local authorities allocate adequate resources and attention to reduce access barriers [37].

Improved human resources are one key element identified as necessary to effectively manage the epidemic, contributing to increased vaccination coverage and improved access to primary healthcare among vulnerable groups in a timely manner. The findings are similar to those highlighted by Sheffel et al.,[38], indicated that instead of concentrating only on increasing personnel numbers, the majority of SSA nations may need to make broader investments in the deployment of the health workforce, enhancements to the health workforce’s performance and capability, and resolving demand restrictions. Strengthen human resources for health by going beyond workforce expansion to include strategic deployment, performance improvement, and capacity building.

Outreach and health promotion services are critical for strengthening the resilience of PHC systems during crises. Outreach activities ensure timely access to communities and deliver targeted health promotion programs, including accessible materials such as Braille for the Deaf and other socially disadvantaged groups. This approach fosters collaboration with key community stakeholders, including local leaders and gatekeepers, to manage epidemics effectively. Community engagement is widely recognized as central to comprehensive and effective pandemic responses, ensuring coordination and cooperation among all relevant stakeholders [39]. Health promotion should take the approach of improved emergency risk communication in braille, audio and sign language interpretation in adapted communication formats to reach out to PWDs in an effective and timely manner [40].

Availability of medical supplies and a reliable transport system are critical elements for building resilience during a crisis, ensuring that essential supplies are delivered to health facilities and communities on time. An efficient transport system strengthens the supply chain for both the procurement and utilization of medical resources. Similar to the funding of Zhang et al., [41] maintaining adequate stockpiles of medical supplies and ensuring functional transport networks before an epidemic is essential for enhancing the resilience of PHC systems in managing pandemics.

Our findings highlight the need for sustained support to exogenous primary healthcare elements that strengthen endogenous system functioning if the PHC system is to be resilient. This is consistent with Behera et al. [42], who identify the four key pillars of primary health care as community participation, intersectoral coordination, appropriate technologies, and supportive mechanisms

Compulsory education systems in sign language interpretation among health workers were identified as a need in order to effectively attend to the deaf. This element ensures that all vulnerable groups are attended to adequately with the best way of interaction with health personnel. This reduces dependence among the deaf, who always seek PHC when there is a caregiver. Consistent with the findings of Martin et al [43] a lack of awareness about the needs of the Deaf persists during medical consultations, and our study indicates that this issue is further exacerbated during crises such as the COVID-19 pandemic. Health workers should receive compulsory training in sign language to ensure the Deaf community can access PHC independently. This reduces reliance on caregivers and addresses gaps in awareness that are worsened during crises like the COVID-19 pandemic [44].

Political will and the Constituency Development Fund (CDF) were identified as key factors in enhancing vaccination coverage and access to primary healthcare during crises. Political commitment supports the financing of PHC services at the community level in Zambia, including direct investments through the CDF aimed at strengthening health infrastructure, improving roads to health facilities, and expanding electrification. The CDF has proven to be an effective mechanism for delivering community-based projects, directly financing PHC development, and addressing the specific needs of constituencies [45]. Although this funding mechanism reflects strong political will, its direct financing of community development, engagement of local populations in decision-making, and responsiveness to concrete community needs can significantly improve vaccination coverage and enhance access to PHC among vulnerable groups [46]. Engage local populations in planning and implementation to enhance ownership, accountability, and effectiveness of health interventions.

According to the findings, there is need for the inclusion of the PWDs in the national insurance scheme to create a resilient primary health care system that is inclusive. Enhancing health insurance aims to achieve various beneficial goals, such as expanding access to healthcare services, minimizing the risk of catastrophic health costs, and bettering health outcomes [47]. This aligns with evidence from other low- and middle-income countries, where health insurance coverage has been associated with increased healthcare utilisation and reduced out-of-pocket expenditures among vulnerable populations [48]. The expressed desire by participants to be included in NHIMA underscores the unmet needs in the current system and the importance of deliberate policy measures to integrate PWDs into national health financing mechanisms.

### Study Strength

Despite the limitations noted, this study has several notable strengths. It is grounded in a disability-inclusive health systems perspective, ensuring that the experiences of persons with disabilities are central to the analysis. The use of participatory systems thinking strengthens the study by enabling the co-production of knowledge with stakeholders and capturing complex interrelationships within the health system. In addition, the active engagement of community stakeholders enhances the credibility, contextual relevance, and practical applicability of the findings. The study is also highly relevant to ongoing global and regional priorities, particularly pandemic preparedness and the pursuit of Universal Health Coverage (UHC). Finally, it makes an important contribution to the growing body of evidence on primary healthcare systems in Africa, particularly in relation to equity and inclusion.

### Study Limitations

This study has several limitations. First, the findings are based on primary data collected from specific communities in Zambia, which may limit the generalizability of the results to other regions or countries. Second, some responses may have been influenced by social desirability bias, particularly when participants discussed sensitive issues related to healthcare access and vulnerability. Thirdly, while the study identified key elements affecting PHC resilience and vaccination coverage, it did not quantify the relative contribution of each factor, which could be explored in future research. Despite these limitations, the study provides valuable insights into strengthening PHC systems and informing policies for vulnerable populations.

## Conclusion

This study demonstrates that resilience in primary healthcare (PHC) systems emerges from the interaction of key system drivers operating through reinforcing and balancing feedback mechanisms, where improved financing strengthens service delivery by enabling the procurement of essential medicines, infrastructure development, and workforce support (reinforcing loop), while inadequacies in funding disbursement create service bottlenecks, staff shortages, and supply interruptions that weaken system performance (balancing loop). Investments in infrastructure, human resources, financing, supply chains, and inclusive service delivery can improve vaccine uptake and access to PHC services. However, these gains are constrained by increasing demand, which places pressure on financial and system capacity. The findings highlight that strengthening transport, outreach, and workforce capacity enhances service accessibility and responsiveness, particularly for PWDs. At the same time, reliable financing mechanism and coordinated community engagement are critical for maintaining continuity during system shocks such as pandemics. Overall, the model underscores the importance of multi-sectoral collaboration and the need to balance resource allocation with rising demand. Addressing these dynamics is essential for sustaining improvements in service delivery, reducing inequities, and advancing progress toward universal health coverage.

## Data Availability

All data produced in the present study are available upon reasonable request to the authors

